# Monitoring COVID-19 progression: Look at Us Today, See Yourself Tomorrow

**DOI:** 10.1101/2020.04.20.20072991

**Authors:** Jungsik Noh

## Abstract

The coronavirus disease 2019 (COVID-19) pandemic is causing public health emergency and economic crisis all over the globe. Being widely spread, the virus can make any place in the world a new epicenter of the possible second wave of outbreaks. To control the pandemic progression, monitoring of the virus spreading is imperative. This paper proposes a simple and robust approach to monitor the COVID-19 pandemic progression in many countries or regions. This data science pipeline can provide actionable insights via straightforward COVID-19 data visualization for many regions at a glance, which informs of relative time delay of the pandemic progression, projected numbers of confirmed cases in the near future, and the sizes of infections.

## Introduction

Severe acute respiratory syndrome coronavirus-2 (SARS-CoV-2) caused the coronavirus disease 2019 (COVID-19) pandemic, which began in Wuhan, China, reportedly on December 12, 2019 (Zhou et al., 2020). The virus hit our entire planet hard, infecting more than 2 million cases and taking lives of more than 140,000, as of April 16, 2020 (Dong et al., 2020). Outside China, the first confirmed case of COVID-19 in each country was reported in Thailand on January 13, 2020, in Japan, January 16, in South Korea, January 20, in the U.S., January 20, and in Taiwan, January 21, according to Wikipedia (retrieved April 16 from COVID-19 webpages of those countries). Since January 22, 2020, the Center for Systems Science and Engineering at Johns Hopkins University has collected and tracked COVID-19 data, which shows that the COVID-19 spread over more than 50 countries before the end of February (Table 1, Supplementary Table 1, retrieved April 16 from Dong et al. (2020)).

**Table 1.** **The dates when the first cases were reported in 50 countries outside China (retrieved April 16, 2020 from Wikipedia and Dong et al. (2020))**.

| ID | Country | date | ID | Country | date |
| --- | --- | --- | --- | --- | --- |
| 1 | Thailand | 1/13/2020 | 26 | Egypt | 2/14/2020 |
| 2 | Japan | 1/16/2020 | 27 | Iran | 2/19/2020 |
| 3 | Korea, South | 1/20/2020 | 28 | Israel | 2/21/2020 |
| 4 | US | 1/20/2020 | 29 | Lebanon | 2/21/2020 |
| 5 | Taiwan | 1/21/2020 | 30 | Afghanistan | 2/24/2020 |
| 6 | Singapore | 1/23/2020 | 31 | Bahrain | 2/24/2020 |
| 7 | Vietnam | 1/23/2020 | 32 | Iraq | 2/24/2020 |
| 8 | France | 1/24/2020 | 33 | Kuwait | 2/24/2020 |
| 9 | Malaysia | 1/25/2020 | 34 | Oman | 2/24/2020 |
| 10 | Nepal | 1/25/2020 | 35 | Algeria | 2/25/2020 |
| 11 | Canada | 1/26/2020 | 36 | Austria | 2/25/2020 |
| 12 | Australia | 1/26/2020 | 37 | Croatia | 2/25/2020 |
| 13 | Cambodia | 1/27/2020 | 38 | Switzerland | 2/25/2020 |
| 14 | Germany | 1/27/2020 | 39 | Brazil | 2/26/2020 |
| 15 | Sri Lanka | 1/27/2020 | 40 | Georgia | 2/26/2020 |
| 16 | Finland | 1/29/2020 | 41 | Greece | 2/26/2020 |
| 17 | United Arab Emirates | 1/29/2020 | 42 | North Macedonia | 2/26/2020 |
| 18 | India | 1/30/2020 | 43 | Norway | 2/26/2020 |
| 19 | Philippines | 1/30/2020 | 44 | Pakistan | 2/26/2020 |
| 20 | Italy | 1/31/2020 | 45 | Romania | 2/26/2020 |
| 21 | Russia | 1/31/2020 | 46 | Denmark | 2/27/2020 |
| 22 | Sweden | 1/31/2020 | 47 | Estonia | 2/27/2020 |
| 23 | United Kingdom | 1/31/2020 | 48 | Netherlands | 2/27/2020 |
| 24 | Spain | 2/1/2020 | 49 | San Marino | 2/27/2020 |
| 25 | Belgium | 2/4/2020 | 50 | Belarus | 2/28/2020 |

To overcome the unprecedented pandemic, accurate and quick monitoring of the virus spreading is critical. Moreover, since the exponentially growing number of confirmed cases and death toll is not intuitive, data science perspective is imperative in controlling the COVID-19 pandemic situation (IHME, 2020; Wu and McGoogan, 2020). A progression of the pandemic outbreak can be visualized by the curve of total confirmed cases or daily new cases over time. Such curves for individual countries show us that some countries like China, South Korea and Italy are at the later stage of the virus spreading, while others might be at the beginning.

Here, a data science pipeline is proposed to monitor and visualize COVID-19 time series. Showing the ever-increasing number of new cases every day, New York Governor in the U.S. sent a message on April 1, 2020, “Look at us today, see yourself tomorrow.” The COVID-19 data visualization presented here shows exactly what the governor said by numbers. By looking at the preceding pandemic trajectories in Italy and South Korea, we may answer how much a country or a region is behind Italy or South Korea on the pandemic progression, so that we can picture how many new cases in the region could be projected in the near future.

An advantage of this model-free approach is robustness against possible dramatic changes in the virus spreading, to which model-based methods can be vulnerable. Using reference countries allows us to compare all the trajectories of the virus spreading in a quantitative way, so it helps monitor the pandemic progression in multiple regions. Because the method uses normalization of population sizes, the normalized trajectory of China was not representative as a reference. Thus, South Korea and Italy were chosen among early infected countries as references, while the pipeline allows any other pandemic trajectories as references.

## Results

To monitor the progression of COVID-19, an informative statistic is the daily growth rate of cumulative confirmed cases. The time courses of the growth rates in many countries showed a pattern (Supplementary Figure 1). The daily growth rates were roughly around 40% with large variability at the beginning, and the rates began to monotonically decrease toward zero after strong restrictive policies to slow down the spreading. As of April 16, 2020, the daily growth rate of total cases in the U.S. is 5.1%, which accounts for 32,076 newly infected and confirmed people, and the growth rate in Italy is 2.3%, according to Wikipedia. It took 16-17 days for Italy’s rates to decrease from ∼5.1% to 2.3%, while the total death toll increased from 11,591 to 22,170 during the period in Italy.

**Figure 1.**
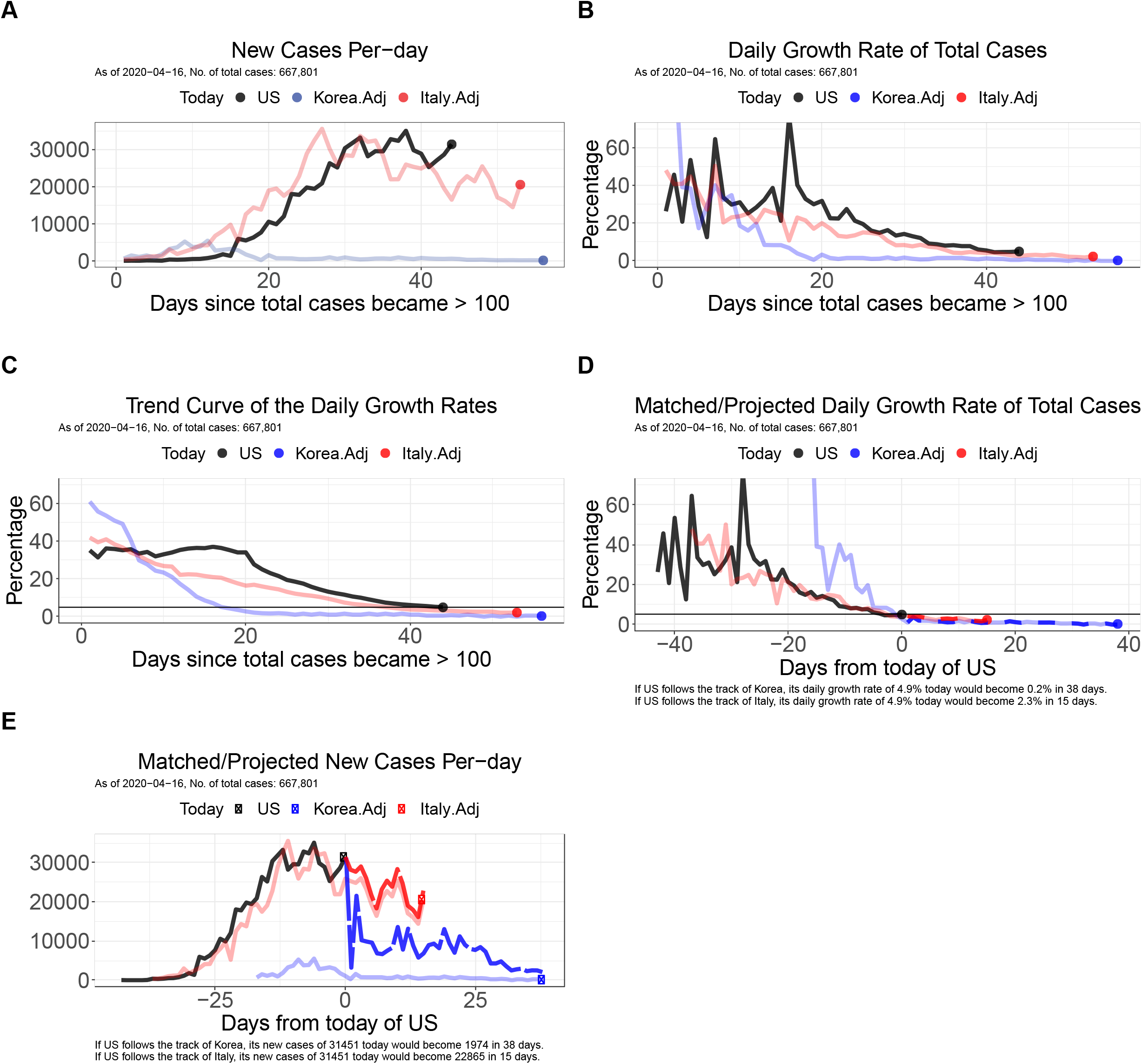
Monitoring COVID-19 time series of daily new confirmed cases and growth rates. **(A)** Daily new confirmed cases in the U.S., South Korea and Italy, after adjusting the population size of South Korea and Italy to the one of the U.S. Each time series starts since total confirmed cases became more than 100 before population adjustment. **(B)** Daily growth rates of total confirmed cases in the U.S., Korea and Italy. Each time series starts since total confirmed cases became more than 100. **(C)** Trend curves of the daily growth rates of total cases for the U.S., Korea and Italy. The value at each day on the trend curves is the average of 9 daily rates around each day, and the most recent value is the average of recent five daily growth rates. **(D)** Three trajectories of daily growth rates of total cases in the U.S., Korea and Italy are shifted in time, so that the daily growth rates of the three countries are close to each other near day = 0. Blue and red dashed lines denote two possible projections for the growth rates of the U.S. obtained from the trajectories of Korea and Italy, respectively. **(E)** Daily new confirmed cases in the U.S., Korea and Italy, after adjusting the population size of Korea and Italy to the one of the U.S. Here, the three trajectories are shifted in time, so that the daily growth rates of the three countries are close to each other near day = 0. Blue and red dashed lines denote two possible projections for the new cases of the U.S. acquired from the growth rates of Korea and Italy, respectively.

The current level of daily growth rate is an informative indicator of the progression stage of COVID-19 pandemic events. In the example of the U.S., the daily growth rate of 4.5% on April 14 became even increased to 5.1% on April 16. This might serve as an early warning in controlling the U.S. pandemic progression. But, current tracking systems of COVID-19 data are incapable of displaying such alarming signals, because they focus on the number of daily new cases, which is not normalized by the growing number of total cases, so fails to expose important changes in the trend of the virus spreading. To monitor the pandemic progression and project the numbers in the coming weeks, it is better to watch the time course of daily growth rates, of which data visualization was not easily available.

Below, with the example of the U.S. that has the most confirmed cases among countries, I present the COVID-19 time series visualization for monitoring the pandemic progression. Figure 1A shows new confirmed cases per-day in the U.S., South Korea and Italy, after adjusting the population size of Italy and South Korea to the one of the U.S. Each time series starts since the total cases became greater than 100 before population adjustment, so that computing daily growth rates of total cases becomes reliable. Figure 1B visualizes daily growth rates of cumulative confirmed cases during this timeline, which are more informative quantities. It should be noted that just a 2 or 3% difference in this daily growth rate has tremendous impact on the total death toll when the differences are accumulated over many days. All the three trajectories of growth rates are decreasing toward zero. Then the level of the rates gives us a hint to where the U.S. is on the COVID-19 trajectories of Italy and South Korea. Figure 3 shows the trend of the daily growth rates in the three countries. The value at each day on the trend curves is the average of 9 daily rates around each day, and the most recent value is the average of recent five daily rates. It is now clearer that the growth rates were roughly in the range of 20–60% at the beginning, and then they are decreasing.

Since the pattern is simple, we may look at the past of Italy or South Korea to see where the pandemic in the U.S. is and would be going. The three trajectories of growth rates in Figure 2 can be shifted in time, so that recent growth rates of the U.S. become the closest to the past growth rates of Italy and Korea at a certain time point. Figure 4 visualizes the three curves in Figure 2 after automatic matching (See Methods). Then, the past trajectories of Italy or Korea from the anchored time point to today give us concrete numbers in each of two possible projections, which the U.S. growth rates of total positive cases might follow from today of the U.S. This simple computation gives us the information about how many days the U.S. trajectory is behind Italy or Korea, and what to expect for the growth rates of the U.S. in the near future.

**Figure 2.**
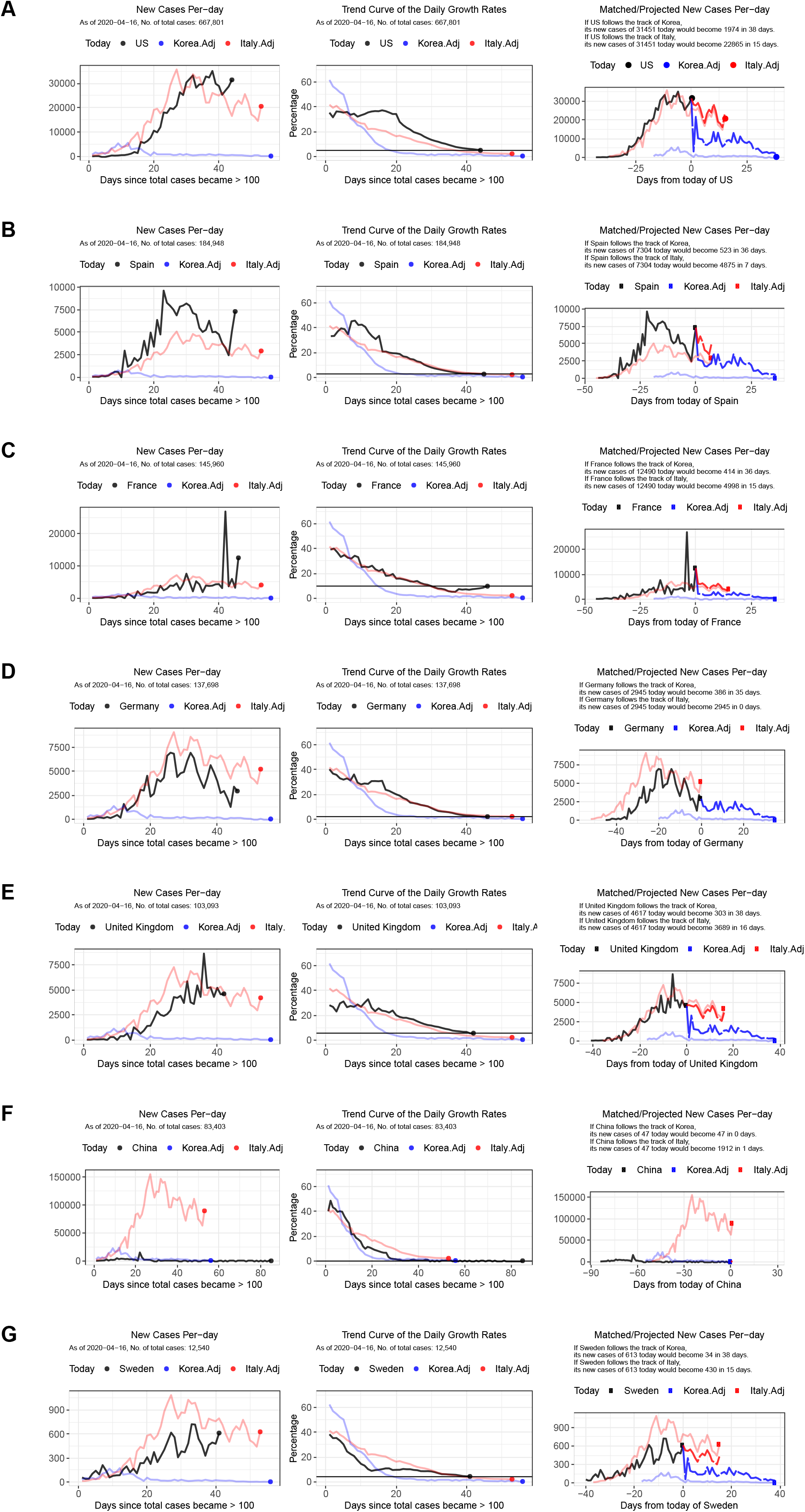
A daily report on the COVID-19 progression in multiple countries. For each country, a daily report lists three plots of new cases per-day, trend curves of the daily growth rates, and the matched/projected new cases per-day, for quick visualization of the COVID-19 progression in multiple countries.

The obtained time differences and projected growth rates can be applied to the current number of total confirmed cases in the U.S., so that we can directly simulate the daily number of total cases in the near future and project the number of new cases (See Methods). Figure 5 shows the matched (time-shifted) and projected time courses of daily new cases of the three countries. Surprisingly, the U.S. curve is shown to be following the Italy curve, at least based on the data up to April 16, 2020. The projection informs us that, as of April 16, if the U.S. follows the track of Korea, its daily new cases of 31,451 today would become 1,974 in 38 days, and that if the U.S. follows the track of Italy, the new cases would become 22,865 in 15 days.

The above COVID-19 time series plots allow us to monitor the pandemic progression in many countries at a glance, potentially providing actionable insights. Figure 2 displays an example of a daily report, which lists three plots of new cases per-day (Figure 1A), trend curves of the daily growth rates (Figure 1C), and the matched/projected new cases per-day (Figure 1E), for each country. The report shows that the daily new cases in Spain dramatically increased after a partial lifting of lockdown restrictions, clearly visualizing slowing down is much harder; the trend of the growth rates of France turned to increasing 8-9 days before, raising an alarming signal; the daily growth rate of Germany is currently close to the one of Italy, so the difference in the pandemic progression between Germany and Italy is estimated as zero day; the U.K. is following the trajectory of Italy 16 days behind. Interestingly, Sweden did not show big differences from other countries in new cases or growth rates, although it did not adopt nation-wide lockdown restrictions.

## Discussion

The proposed data science pipeline of COVID-19 time series visualization will help us monitor the number of confirmed cases and the growth rates every day. It provides straightforward visualization to see where we are in view of the past of Italy and South Korea. Since the analysis compares population-normalized numbers of daily new cases, it also allows us to appreciate the sizes of the infection in different countries or regions. The projected number of daily new cases or growth rates are based on simple calibration and a strong assumption. They are not intended for forecasting, but for monitoring of the current COVID-19 pandemic progression.

The above time series plots for other countries and the states in the U.S. are being updated daily. All the data visualization, daily reports, source codes in R, and projected numbers are openly available at a github repository (https://github.com/JungsikNoh/COVID-19_LookAtUsToday_SeeYourselfTomorrow).

## Data sources

The number of confirmed cases for countries are from COVID-19 data repository of Johns Hopkins CSSE (https://github.com/CSSEGISandData/COVID-1).

## Methods

### Automatic time matching

To match (time-shift) trajectories of the growth rates, I computed the mean squared differences (MSD) between the growth rates of recent three days in the U.S. and the rates of consecutive three days in the reference country, as follows:

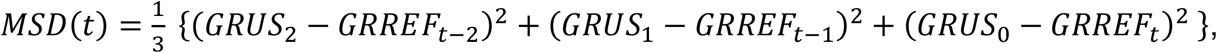

where *GRUS*_*i*_ is the growth rate of the U.S. *i*-day before, and *GRREF*_*t*-1_ is the rate of the reference country at (*t* −i) day. Then, I computed the time t when the *MSD*(*t*) is minimized, and shifted the U.S. trajectory to the past by t days.

### Projecting the number of new cases

Suppose that the current daily growth rate of the U.S. is matched with the rate of the reference country at day *t*_0_ Then, the reference rates after the day, 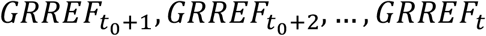, are taken as a potential scenario for the future growth rates of the U.S. Denote by *TUS*_*h*,_ *NCUS*_*h*_ the total confirmed cases and daily new cases of the U.S. *h* days later, respectively. Then, the total cases and new cases can be projected in the near future as follow:

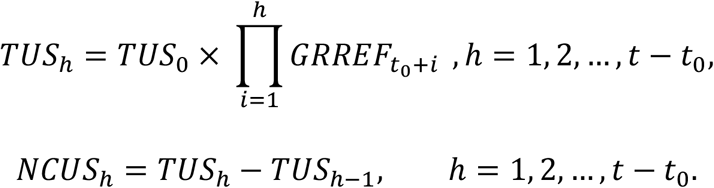

## Data Availability

All data is publicly available.

https://github.com/JungsikNoh/COVID-19_LookAtUsToday_SeeYourselfTomorrow

## Supplementary materials

**Supplementary Figure 1. An animation illustrates that decreasing daily growth rates of total cases follow the preceding trajectory of Italy with certain time lags in five countries undergoing severe COVID-19 outbreaks**.

Each time series starts after total confirmed cases became more than 100. Several numbers of total cases in March, 2020 were manually corrected according to Wikipedia, since the total cases were incorrectly the same as the ones of yesterday.

**Supplementary Table 1. The dates when the first cases were reported in ∼200 countries outside China (retrieved April 16, 2020 from Wikipedia and Dong et al. 2020)**.

